# Humoral Immune Responses in German Dialysis Patients after mRNA Omicron JN.1 Vaccination

**DOI:** 10.1101/2024.09.17.24313789

**Authors:** Metodi V. Stankov, Markus Hoffmann, Christine Happle, Karsten Lürken, Amy Kempf, Inga Nehlmeier, Andrea Stölting, Stefan Pöhlmann, Alexandra Dopfer-Jablonka, Georg M. N. Behrens

## Abstract

To assess the effect of the updated mRNA JN.1 omicron vaccine (bretovameran, BioNTech/Pfizer, Mainz, Germany) in an immunocompromised and elderly population, we measured humoral immune responses after mRNA omicron JN.1 vaccination in 37 haemodialysis patients before and 21 days after vaccination.

We observed a 3-fold change in anti-S IgG, and a 4·7-fold change in anti-S omicron IgG. Memory B cells (MBC) exclusively binding the receptor binding domain (RBD) of JN.1 displayed a median frequency of 0·11% before vaccination and changed significantly 3·9-fold to a median of 0·43%. Cross reactive JN.1 RBD and Wuhan-Hu-1 S-binding MBCs and MBCs only binding to Wuhan-Hu-1 S changed 2·3-fold and 1·8-fold, respectively. Using a vesicular stomatitis virus-based pseudovirus particle (pp) neutralisation assay, baseline response rates were 86% for XBB.1.5_pp_, 78% for JN.1_pp_, 73% for and KP.2_pp_, 65% for KP.2.3_pp_ and KP.3_pp_, and 68% for LB.1_pp_. After vaccination, the response rates for all pseudoviruses increased significantly, and we observed a mean increase in neutralisation of XBB.1.5_pp_, JN.1_pp_, KP.2_pp_, KP.2.3_pp_, KP.3_pp_, and LB.1_pp_ of 8·3-fold, 18·7-fold, 22·5-fold, 18·7-fold, 25·5-fold, and 23·5-fold, respectively. In summary, our report provides first evidence for a firm humoral immune response in dialysis patients after mRNA omicron JN.1 vaccination.

Our data suggest that the vaccine could be highly effective at enhancing protection of vulnerable populations against evolving SARS-CoV-2 variants.

---

We recently demonstrated the impact of an updated, JN.1 omicron sublineage adapted SARS-CoV-2 mRNA vaccine on neutralising antibodies in healthy individuals [1]. To assess the effect of this updated vaccine in an immunocompromised and elderly population, we measured humoral immune responses after mRNA omicron JN.1 vaccination in haemodialysis patients. Patients undergoing haemodialysis display reduced and less sustainable immune responses after vaccination as compared to healthy individuals [2] and exhibit an elevated risk for severe COVID-19 [3], including upon infection with the omicron variant [4]. Here, we quantified changes in JN.1-reactive memory B cells (MBC) and neutralising antibody titres against contemporary omicron variants in 37 dialysis patients who received 30 μg of the updated mRNA omicron JN.1 vaccine (bretovameran, BioNTech/Pfizer, Mainz, Germany). The median age of patients was 68 years (range 28–90 years), 26 (70%) were male, 16 (43%) reported previous SARS-CoV-2 infections, and 37 (100%) had received previous vaccinations against SARS-CoV-2 (appendix p 2).

Median pre-vaccination anti-spike (anti-S) IgG antibody levels were 1817·0 binding antibody units (BAU) per mL (Interquartile Range (IQR) 2670·9), and median anti-omicron spike IgG levels were 170·4 relative units (RU) per mL (IQR 266·1). Twenty-one days after vaccination, we observed a 3-fold change to a median of 5413·0 BAU per mL in anti-S IgG (IQR 9024·0), and a 4·7-fold change to a median of 796·0 RU per mL in anti-S omicron IgG (IQR 758·8; figure A).

**Figure:**
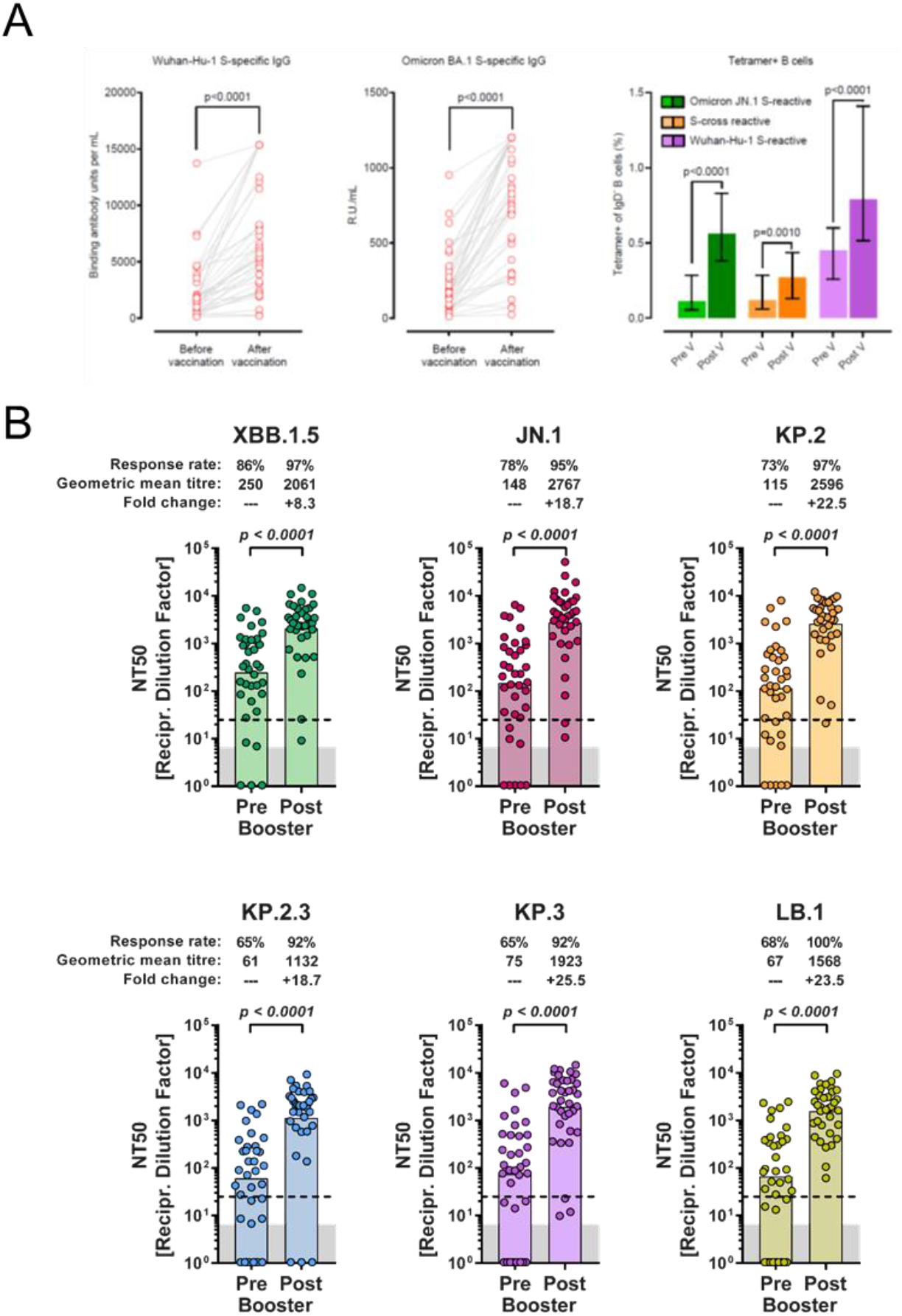
Humoral immune responses in haemodialysis patients following mRNA omicron JN.1 vaccination. (A) Concentrations of Wuhan-Hu-1 S-reactive IgG and omicron BA.1 S-reactive IgG in plasma (n=37) obtained before or after vaccination with the mRNA omicron JN.1 vaccine (*left and middle panels*). Frequencies of MBC (n=37) binding the receptor-binding domain of JN.1 (green), to Wuhan-Hu-1 spike (pink), or both (cross-reactive, orange) before and after JN.1 vaccination *(right panel)*. Data are represented as median (coloured bars) + IQR (whiskers). Statistical significance was assessed by Wilcoxon matched-pairs signed rank test. (B) Neutralisation of vesicular stomatitis virus-based pseudovirus particles harboring the indicated S proteins by donor-matched plasma (n=37) taken before or after vaccination with the mRNA omicron JN.1 vaccine. Data represent GMT (colored columns) from a single experiment, conducted with four technical replicates. The lowest plasma dilution tested (dashed lines) and the assay threshold (lower limit of detection; grey shaded areas) are indicated. Information on response rates and mean fold change in neutralisation after vaccination are indicated above the graphs. Statistical significance was assessed by Wilcoxon matched-pairs signed rank test. Of note, for graphical reasons, plasma samples yielding a neutralization titre 50 (NT50) value below 6·25 (limit of detection) were manually placed at the bottom of the axis. Individual neutralisation data are available in the appendix (pp 11-12). IQR=interquartile range. NA=not applicable. S=spike. GMT=geometric mean titres.

The proportion of MBCs binding SARS-CoV-2 S increased following JN.1 vaccination. Specifically, MBCs exclusively binding the receptor binding domain (RBD) of JN.1 displayed a median frequency of 0·11% before vaccination and changed significantly 3.9-fold to a median of 0·43%. Cross reactive JN.1 RBD and Wuhan-Hu-1 S-binding MBCs and MBCs only binding to Wuhan-Hu-1 S changed 2·3-fold and 1·8-fold, respectively (figure A, appendix p 8-9).

Next, we assessed changes in plasma neutralisation upon vaccination with the updated JN.1 omicron mRNA vaccine. For this, we utilized a vesicular stomatitis virus-based pseudovirus particle (pp) neutralisation assay. Specifically, we used pseudoviruses harboring S proteins of six SARS-CoV-2 lineages (figure B, appendix pp 10-12). Before JN.1 vaccination, baseline response rates were 86% for XBB.1.5_pp_, 78% for JN.1_pp_, 73% for and KP.2_pp_, 65% for KP.2.3_pp_ and KP.3_pp_, and 68% for LB.1_pp_ (figure B). Further, particles bearing KP sublineage or LB.1 S proteins were less efficiently neutralised compared with JN.1_pp_ (mean change, 1·3-fold to 2·4-fold), indicating antibody evasion (appendix p10). After vaccination, the response rates for all pseudoviruses increased significantly, and we observed a mean increase in neutralisation of XBB.1.5_pp_, JN.1_pp_, KP.2_pp_, KP.2.3_pp_, KP.3_pp_, and LB.1_pp_ of 8·3-fold, 18·7-fold, 22·5-fold, 18·7-fold, 25·5-fold, and 23·5-fold, respectively (figure B, appendix p 10).

In summary, our report provides first evidence for a firm humoral immune response in dialysis patients after mRNA omicron JN.1 vaccination. The triple increase in MBCs reacting with the RBD of omicron JN.1 S combined with the strongly amplified neutralisation against JN.1 and other contemporary omicron variants suggests that the updated mRNA omicron JN.1 vaccine could be highly effective at enhancing protection of vulnerable populations [3,4] against evolving SARS-CoV-2 variants.

## Data Availability

All requests for raw and analysed data that underly the results reported in this article will be reviewed within four weeks by the CoCo Study Team, Hannover Medical School to determine whether the request is subject to confidentiality and data protection obligations. Data that can be shared will be released via a material transfer agreement.

## Appendix

### Supplementary Table

**Supplementary Table S1.**
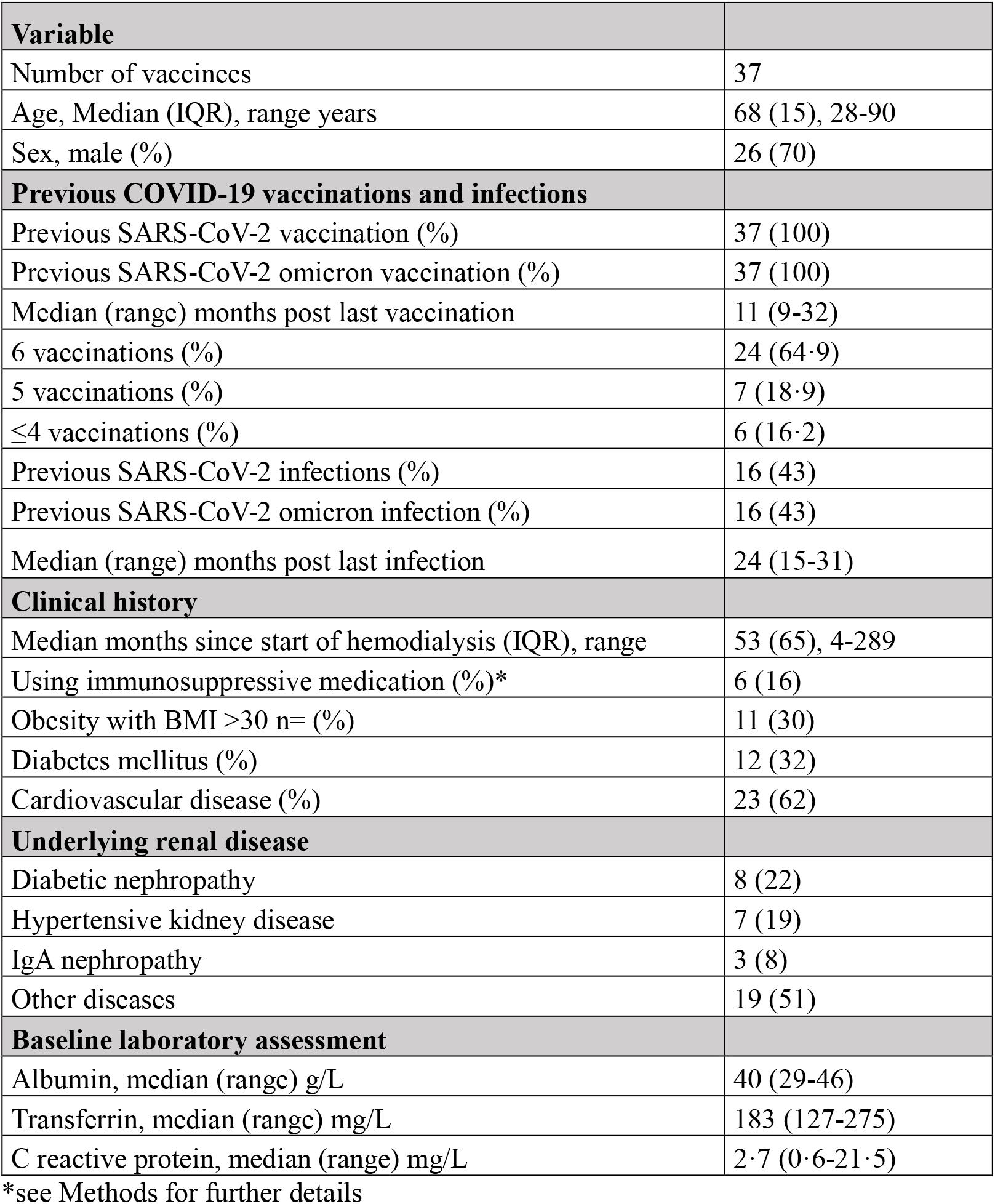
Demographics, vaccination and disease history of included dialysis patients.

| **Variable** |  |
| --- | --- |
| Number of vaccinees | 37 |
| Age, Median (IQR), range years | 68 (15), 28-90 |
| Sex, male (%) | 26 (70) |
| **Previous COVID-19 vaccinations and infections** |  |
| Previous SARS-CoV-2 vaccination (%) | 37 (100) |
| Previous SARS-CoV-2 omicron vaccination (%) | 37 (100) |
| Median (range) months post last vaccination | 11 (9-32) |
| 6 vaccinations (%) | 24 (64·9) |
| 5 vaccinations (%) | 7 (18·9) |
| ≤4 vaccinations (%) | 6 (16·2) |
| Previous SARS-CoV-2 infections (%) | 16 (43) |
| Previous SARS-CoV-2 omicron infection (%) | 16 (43) |
| Median (range) months post last infection | 24 (15-31) |
| **Clinical history** |  |
| Median months since start of hemodialysis (IQR), range | 53 (65), 4-289 |
| Using immunosuppressive medication (%)* | 6 (16) |
| Obesity with BMI >30 n= (%) | 11 (30) |
| Diabetes mellitus (%) | 12 (32) |
| Cardiovascular disease (%) | 23 (62) |
| **Underlying renal disease** |  |
| Diabetic nephropathy | 8 (22) |
| Hypertensive kidney disease | 7 (19) |
| IgA nephropathy | 3 (8) |
| Other diseases | 19 (51) |
| **Baseline laboratory assessment** |  |
| Albumin, median (range) g/L | 40 (29-46) |
| Transferrin, median (range) mg/L | 183 (127-275) |
| C reactive protein, median (range) mg/L | 2·7 (0·6-21·5) |

\*see Methods for further details

## Methods

### Participants

Haemodialysis patients were recruited as part of the COVID-19 Contact (CoCo) Study (German Clinical Trial Registry, DRKS00021152). This is a prospective, observational study monitoring anti-SARS-CoV-2 immune responses in healthcare professionals and patients [1]. Initially, n=52 dialysis patients were immunized with 30μg of JN.1 Comirnaty^®^ omicron/Bretovameran. Blood was drawn shortly before and 21 days after vaccination. Eight patients were excluded from the analysis due to confirmed SARS-CoV-2 infections during the observational period after vaccination, and n=7 patients were lost to follow up at day 21 due to death, hospitalization, or other (unspecified) reasons. We finally analysed immune responses in n=37 dialysis patients. Details on the demographic data and case history of the included patients can be found in Suppl. Table 1. In summary, all patients underwent hemodialysis, and the median duration of dialysis until JN.1 vaccination had been 53 months (range 4-289 months). Most frequent underlying renal diseases were diabetic nephropathy (22%), followed by hypertensive kidney disease (19%), and IgA nephropathy (8%). Thirty percent of patients were obese, 32% suffered from diabetes mellitus, and 62% reported underlying cardiovascular disease. Six patients (16%) received immunosuppressive medication. Four of them received treatment with oral corticosteroids (5mg prednisolone in three and 20 mg hydrocortisone in one), one patient received a combination of Tacrolimus and Everolimus, and one further patient was under treatment with Daratumumab.

Sample size calculations estimated that a sample size of n=43 should be sufficient for detection of a clinically relevant difference within the group, assuming that mean SARS-CoV-2 S1 protein-specific IgG levels of 2385 (SD=2394) BAU/ml double from pre-vaccination levels. Power calculation was performed using G*Power, Version 3.1.9.6. and based on anti-S IgG analyses in our previously published cohort of n=44 patients from previous recruitment phase of the CoCo dialysis study before XBB.1.5 vaccination [7], which is our best available estimate of pre-vaccination levels, correlation between groups 0.5. The estimate was based on a 2-tailed paired t-test of mean differences, with 98% power and 5% significance level. Based on our previous work, a loss-to follow-up rate of 20% was estimated. Following these calculations, we aimed at a sample size of n=52 vaccinated hemodialysis patients.

### Serology

Serology was performed as previously described [1]. Briefly, we measured SARS-CoV-2 IgG by quantitative ELISA (Anti-SARS-CoV-2 QuantiVac-ELISA, EI 2606-9601-10G, and Anti-SARS-CoV-2 Omikron-ELISA, EI 2606-9601-30 G, both EUROIMMUN, Lübeck, Germany) according to the manufacturer’s instructions (dilution up to 1:4,000). We used anti-S concentrations expressed as relative units (RU)/mL as assessed from a calibration curve with values above 11 RU/mL defined as positive. Values above the upper quantification limit of the assay (120 RU/mL for a 1:100 dilution) are set to 120 RU/mL (adapted to the used dilution) and used for further analysis.. We provide results obtained with the QuantiVac ELISA in binding antibody units (BAU/mL), which were converted by multiplying RU/mL by 3·2, as specified by the manufacturer. We performed anti-SARS-CoV-2 nucleocapsid (NCP) IgG measurements according to the manufacturer’s instructions (EUROIMMUN, Lübeck, Germany). We used an AESKU.READER (AESKU.GROUP, Wendelsheim, Germany) and the Gen5 2.01 Software for analysis.

### Flow cytometric detection and analysis of SARS-CoV-2-specific B cells

For tetramer preparation, we used recombinant, biotinylated SARS-CoV-2 S protein (Wuhan-Hu-1 and omicron/JN.1) to detect SARS-CoV-2-spike-reactive B cells. Tetramerisation was performed as previously described [1]. Briefly, we tetramerised recombinant Wuhan-Hu-1 S proteins with fluorescently labelled streptavidin/R-phycoerythrin conjugate (Cat# S21388, ThermoFisher) and recombinant spike (RBD) omicron/JN.1 protein with fluorescently labelled streptavidin/ allophycocyanin (Cat# S868; ThermoFisher) [1]. We isolated fresh PBMCs samples, washed and re-suspended them in FACS buffer (PBS, 1 mg/mL BSA, 1 mmol/L EDTA) and stained cells with antibodies (Table S3) and tetramerised recombinant proteins against the Wuhan-Hu-1 S and omicron JN.1 S (RBD) for 20 min at room temperature. After two more wash steps, we acquired samples on a spectral flow cytometer (Cytek Northern Lights) and analysed data using SpectroFlo and/or FCS Express software according to the gating strategy (Figure S2).

### Production of vesicular stomatitis virus-based pseudovirus particles and pseudovirus neutralisation test (pVNT)

pVNTs were performed according to a previously published protocol [2]. In brief, 293T cells were transfected with S protein expression plasmid. Expression plasmids pCG1_SARS-2-SΔ18 XBB.1.5 (EPI_ISL_16239158; codon-optimised, deletion of last 18 aa residues at the C-terminus) [3] and pCG1_SARS-2-SΔ18 JN.1 (EPI_ISL_18530042; codon-optimised, deletion of the last 18 aa residues at the C-terminus) [4] have been described before. In addition, S protein expression plasmids pCG1_SARS-2-SΔ18 KP.2 (EPI_ISL_19197864; codon-optimised, deletion of the last 18 aa residues at the C-terminus), pCG1_SARS-2-SΔ18 KP.2.3 (EPI_ISL_19197559; codon-optimised, deletion of the last 18 aa residues at the C-terminus), pCG1_SARS-2-SΔ18 KP.3 (EPI_ISL_19203001; codon-optimised, deletion of last 18 aa residues at the C-terminus), and pCG1_SARS-2-SΔ18 LB.1 (EPI_ISL_19067004; codon-optimised, deletion of the last 18 aa residues at the C-terminus) were generated by introduction of the required mutations into plasmid pCG1_SARS-2-SΔ18 JN.1 (Figure S1). This was achieved by overlap-extension PCR using overlapping primers that harbour the respective mutations. At 24h posttransfection, the 293T cells were inoculated with a replication-deficient VSV vector that lacks the genetic information for the VSV glycoprotein and instead encodes for an enhanced green fluorescent protein and a firefly luciferase, VSV*ΔG-FLuc (kindly provided by Gert Zimmer, Institute of Virology and Immunology, Mittelhäusern, Switzerland) [5]. Following 1h of incubation at 37 °C and 5% CO_2_, the cells were washed with PBS and further incubated with medium containing anti-VSV-G antibody (culture supernatant from I1-hybridoma cells; ATCC no. CRL-2700) to neutralise residual input virus. After 16-18h of incubation at 37 °C and 5% CO_2_, the pseudovirus-containing supernatant was harvested, centrifuged (4,000 x g, 10 min) to remove cellular debris, and clarified supernatants were stored at -80 °C until further use.

For pVNTs, Vero76 cells (kindly provided by Andrea Maisner, Institute for Virology, Phillips University Marburg) were grown to confluence in 96-well plates. Next, pseudovirus particles were mixed with serial dilutions of heat-inactivated (56 °C, 30 min) plasma, incubated for 30 min at 37 °C, and finally inoculated onto the Vero cells in four technical replicates. Plasma dilutions were prepared in culture medium (final dilution range 1:25 to 1:6,400) and pseudovirus particles mixed with medium without plasma sample served as reference. At 16-18h postinoculation, pseudovirus infection was analysed. For this, the culture supernatant was removed and cells were lysed with PBS containing 0·5 % Tergitol (Carl Roth; 30 min at room temperature). Thereafter, the cell lysates were transferred into white 96-well plates, mixed with luciferase substrate (Beetle-Juice, PJK) and incubated for 1 min, before luminescence was recorded using a Hidex Sense Microplate Reader Software (version 0.5.41.0). Efficiency of neutralisation was determined based on the relative inhibition of pseudovirus infection of Vero76 cells. Signals obtained for cells infected with pseudovirus particles incubated in the absence of plasma served as reference (no inhibition). Next, a non-linear regression model was used to calculate the neutralising titre 50 (NT50), which indicates the plasma dilution responsible for half-maximal inhibition. Of note, plasma samples that yielded NT50 values below 25 (lowest dilution tested) were defined as non-responders and samples that yielded NT50 values below 6·25 (limit of detection, LOD) were assigned an NT50 value of 3·125 (0.5 of LOD).

### Statistics

GraphPad Prism 8.4 or 9.0 (GraphPad Software, USA) and SPSS 20.0.0 (IBM SPSS Statistics, USA) were employed for the statistical analyses. We included outliers into the analysis and excluded missing values pairwise. Mean (SD) was reported for normally distributed data, and median (IQR) for non-normally distributed values. For assessing differences within groups in non-normally distributed data, Wilcoxon matched-pairs signed rank test were employed. We transformed neutralisation titres to geometric mean titres before further analysing this data.

**Fig. S1.**
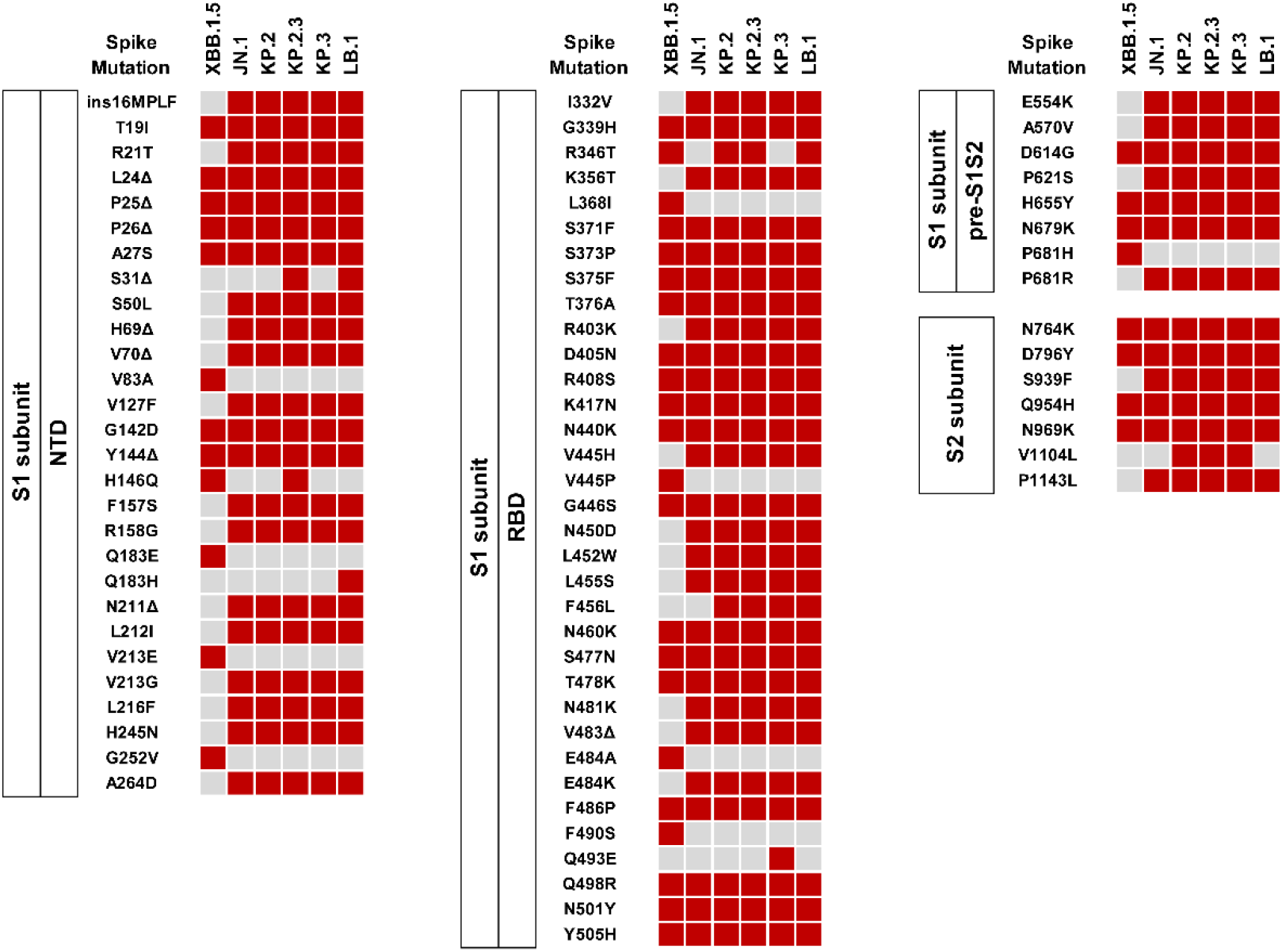
Overview of SARS-CoV-2 lineage-specific spike protein mutations.

**Table S2.** Tetramer Antigen Conjugate.

| Antigen | Conjugate | Clone | Order no. | Company | Dilution |
| --- | --- | --- | --- | --- | --- |
| SARS-CoV-2 Spike Trimer (Wuhan Hu-1) | Biotinylated | NA | #SPN-C82E9 | Acrobiosystems | 100 |
| SARS-CoV-2 Spike Trimer (Omicron RBD JN.1) | Biotinylated | NA | #0BB1D-F001 | Acrobiosystems | 400 |

**Table S3.** Antibody panel.

| Antigen | Conjugate | Clone | Order no. | Company | Dilution |
| --- | --- | --- | --- | --- | --- |
| CD16 | Alexa Fluor 488 | 3G8 | 302019 | BioLegend | 100 |
| CD14 | PE/Cy5 | M5E2 | 301864 | BioLegend | 100 |
| CD20 | Brilliant Violet 570 | 2H7 | 302332 | BioLegend | 100 |
| CD38 | APC-Fire810 | HIT2 | 303550 | BioLegend | 100 |
| CD3 | Alexa Fluor 532 | UCHT1 | 58-0038-42 | Invitrogen | 100 |
| CD27 | Alexa Fluor 700 | O323 | 56-0279-42 | Invitrogen | 100 |
| CD19 | Pacific Blue | SJ25C1 | 363036 | BioLegend | 100 |
| IgD | BV480 | 1A6-2 | 566138 | BD Horizon | 100 |
| Viability | Zombie NIR | NA | 423106 | BioLegend | 1000 |

**Fig. S2.**
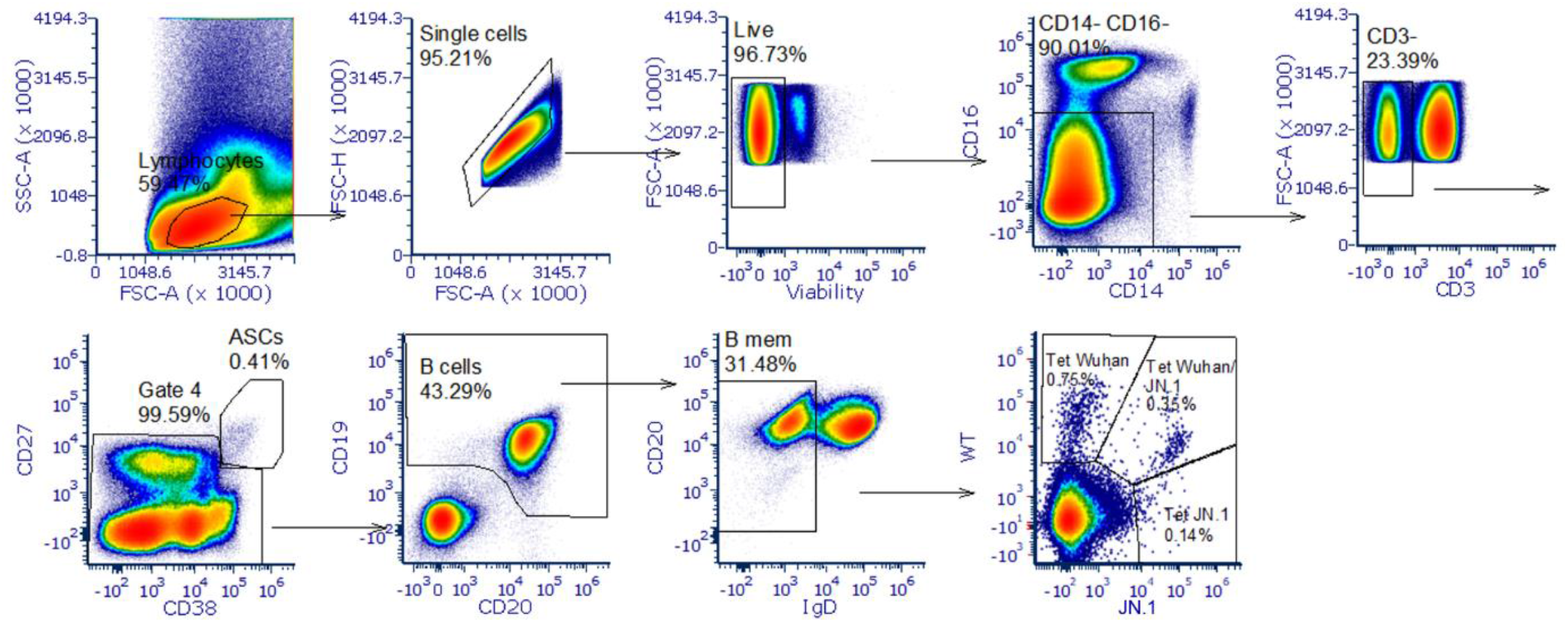
Gating strategy for SARS-CoV-2 S-reactive IgD^-^ cell populations in peripheral blood mononuclear cells. Tetramerised recombinant spike proteins from Wuhan Hu-1 (Tet Wuhan) or the receptor binding domain of spike omicron JN.1 (Tet JN.1) variants were used to identify memory B cells carrying B cell receptors binding to either one or both spike proteins. Pseudocolor plots show representative data from one individual. ASC, antibody secreting cells.

## Limitations of the study and considerations for data interpretation

Our study has some limitations. Our data can only provide first insights into the immune response to the updated JN.1 vaccine in an immunocompromised population. Further data is necessary to assess immune trends, sustainability, and clinical relevance of our findings. SARS-CoV-2 neutralisation was assessed by pVNT, which has been shown to serve as an adequate surrogate model for this purpose [5]. Post vaccination titres in some individuals were at the upper range of the dilutions used. Our findings await confirmation with clinical isolates and eventually validation in clinical studies. Pre-vaccination neutralisation and anti-S IgG responses in haemodialysis patients were lower than previously reported for health care-workers [6]. Whilst median time since last vaccination was similar and haemodialysis patients had received on average one more SARS-CoV-2 vaccination, the rate of hybrid immunity in these patients was 50% less, they were on average 20 years older, had more co-morbidities, and their blood was collected at day 21 (instead of day 13). We believe that these factors are likely influencing the magnitude of the post-vaccination response and limit direct comparisons. Finally, we wish to point out that in the current study we employed tetramers for MBC staining, which bind to the RBD of omicron JN.1 only. This is relevant when comparing changes in different antigen-specific MBC compartments to our previous study in haemodialysis patients after omicon XBB.1.5 vaccination [7], in which we used tetramers reacting to XBB.1.5 spike protein.

## Contributions

Study design: G.M.N.B., A.D.-J.

Data collection and curation: C.H., K.L., A.S.

Investigation: A.K., I.N., M.V.S., M.H.

Data analysis: G.M.N.B, A.D.J., C.H., M.H., M.V.S.

Data interpretation: M.H., S.P., M.V.S., G.M.N.B., A.D.-J.

Writing: C.H., G.M.N.B. with comments from all authors.

G.M.N.B and A.D.-J. have directly accessed and verified the underlying data reported in the manuscript.

## Ethics committee approval

The CoCo Study (German Clinical Trial Register DRKS00021152) and all analyses conducted for this article were approved by the Internal Review Board of Hannover Medical School (institutional review board no. 8973_BO_K_2020, last amendment Aug 2024). All study participants gave written informed consent and received no compensation.

## Funding

A.D.-J. and G.M.N.B. received funding (Niedersächsisches Ministerium für Wissenschaft und Kultur; 14-76103-184, COFONI Network, project 4LZF23), G.M.N.B. acknowledges funding by the European Regional Development Fund ZW7-85151373, and A.D.-J. acknowledges funding by European Social Fund (ZAM5-87006761). S.P. received funding by the EU project UNDINE (grant agreement number 101057100), the COVID-19-Research Network Lower Saxony (COFONI) through funding from the Ministry of Science and Culture of Lower Saxony in Germany (14-76103-184, projects 7FF22, 6FF22, 10FF22) and the German Research Foundation (Deutsche Forschungsgemeinschaft, DFG; PO 716/11-1). No funding sources had any role in the design and execution of the study, writing of the manuscript and/or decision to submit the manuscript for publication. None of the authors received payment by a pharmaceutical company or other agency to write the publication. The authors were not precluded from accessing data in the study, and they accept responsibility to submit for publication.

We thank all patients for their participation, all CoCo study team members and the team of the Dialysis Center Eickenhof for their support. We would like to particularly thank Simon F. Ritter, Louis Kuhnke, Eva Stöppelmann, Melanie Ignacio, Marion Hitzigrath, Noemä Calderón Hampel, Anna-Sophie Moldenhauer, and Luise Graichen for their technical and logistical support.

---

**Table. S4.** Anti-S IgG and anti-S reactive MBC pre and post mRNA JN.1 vaccination immune responses following mRNA omicron JN.1 vaccination.

|  | Median | IQR |
| --- | --- | --- |
| Pre-Vac Wuhan-Hu-1 S-specific IgG [BAU/mL] | 1817 | 2670·9 |
| Post-Vac Wuhan-Hu-1 S-specific IgG [BAU/mL] | 5413 | 9024 |
| Pre-Vac Omicron S-specific IgG [RU/mL] | 170 | 266·1 |
| Post-Vac Omicron S-specific IgG [RU/mL] | 796 | 758·8 |
| Pre-Vac Wuhan-Hu-1 S-reactive MBC (%) | 0·45 | 0·34 |
| Post-Vac Wuhan-Hu-1 S-reactive MBC (%) | 0·79 | 0·90 |
| Pre-Vac cross-reactive MBC (%) | 0·12 | 0·23 |
| Post-Vac cross-reactive MBC (%) | 0·27 | 0·31 |
| Pre-Vac omicron JN.1 RBD-reactive MBC (%) | 0·11 | 0·23 |
| Post-Vac omicron JN.1 RBD-reactive MBC (%) | 0·43 | 0·50 |

**Fig. S3.**
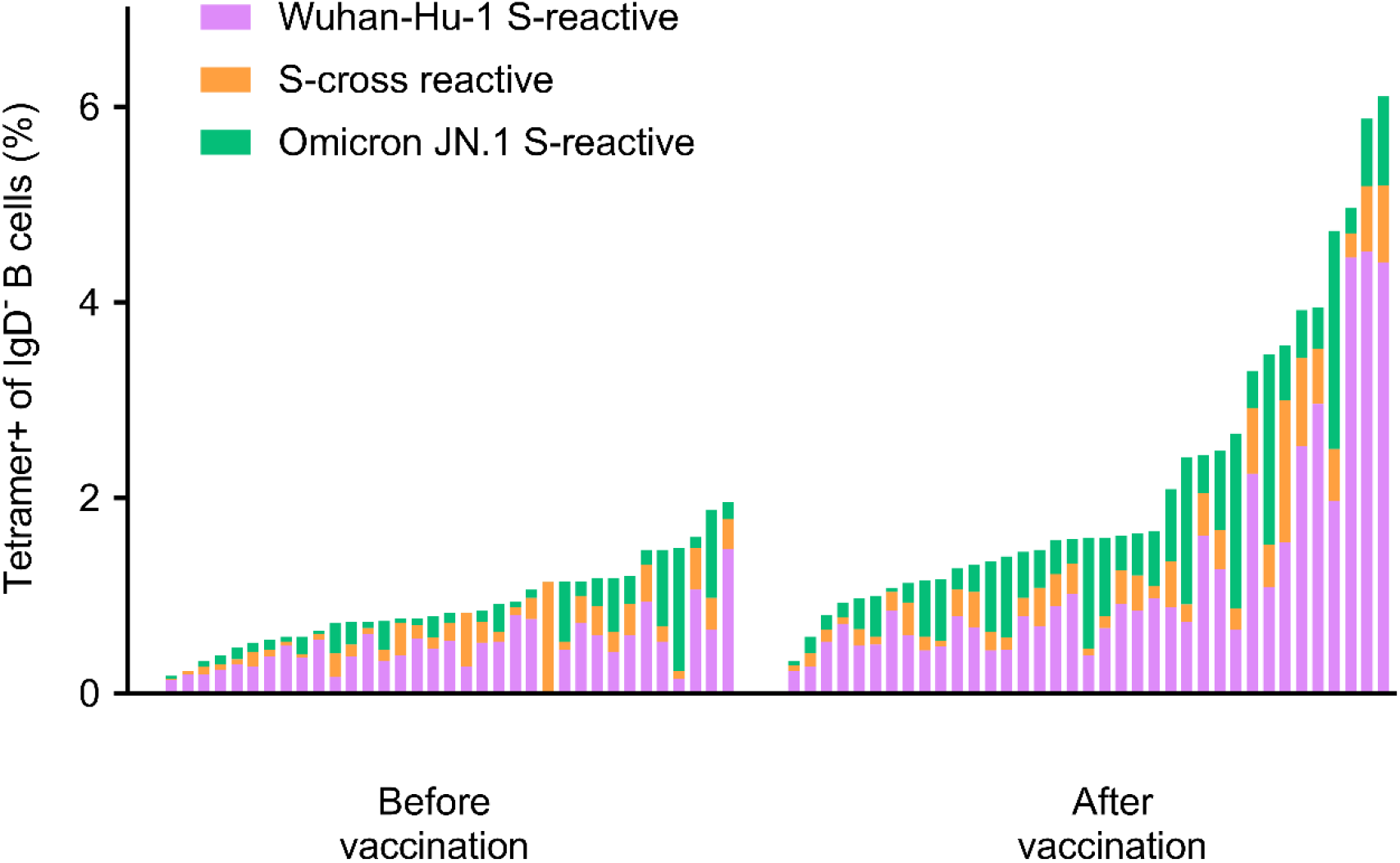
Individual data of changes in SARS-CoV-2 S-reactive IgD^-^ cell populations in peripheral blood mononuclear cells before and post mRNA omicron JN.1 vaccination. Tetramerised recombinant spike proteins from Wuhan-Hu-1 and the receptor binding domain of spike omicron JN.1 (Tet JN.1) variants were used to identify memory B cells carrying B cell receptors binding to either one or both spike proteins (S-cross reactive). B cells gated as shown in Fig. S2.

**Fig. S4.**
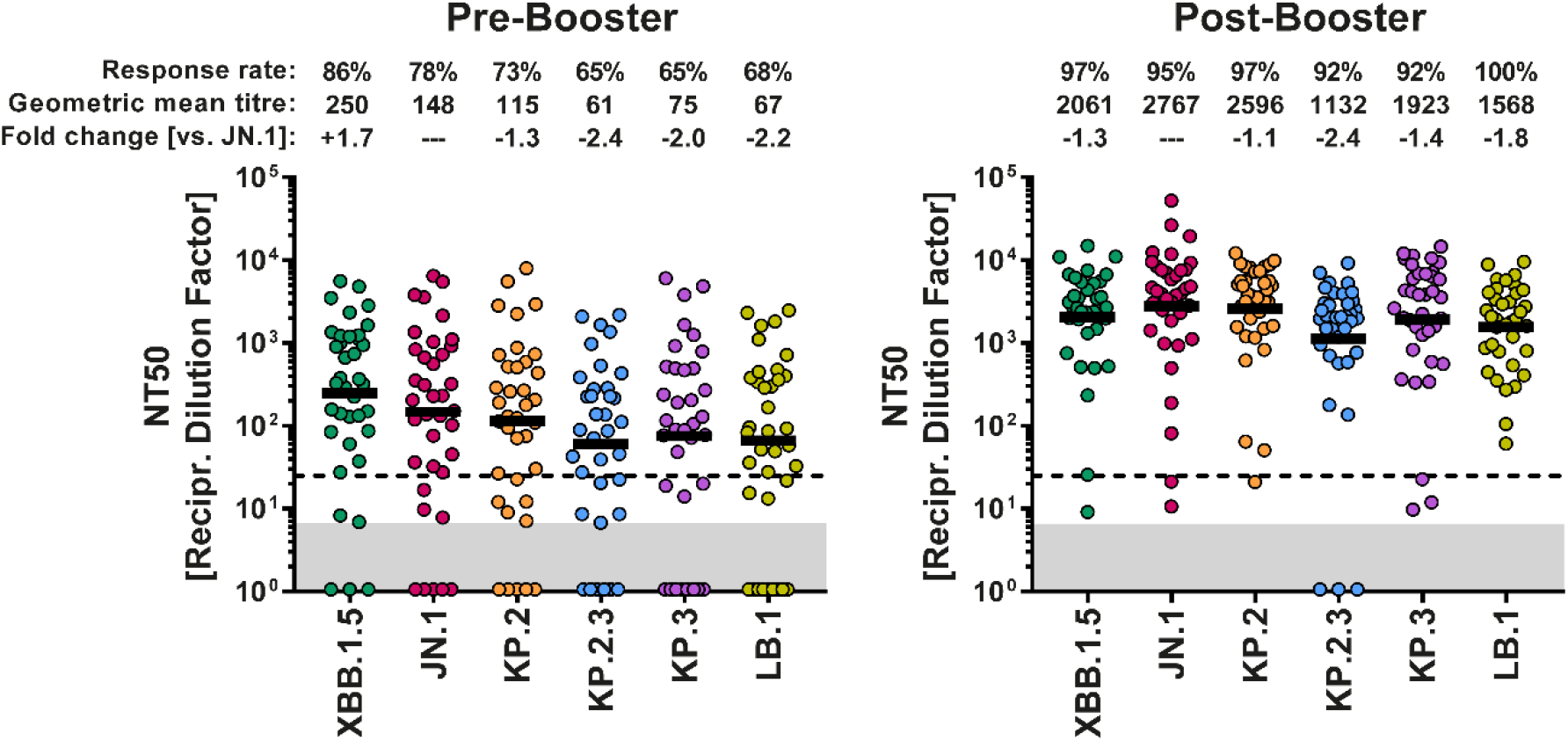
Humoral immune responses following mRNA omicron JN.1 vaccination. The data presented in this panel were regrouped from figure B to compare differences in SARS-CoV-2 lineage-specific neutralisation at baseline (before vaccination) and after vaccination. Information on GMT (also indicated by horizontal lines) response rates, and mean fold change in neutralisation compared with JN.1 pseudovirus particles are indicated above the graphs.

**Fig. S5.**
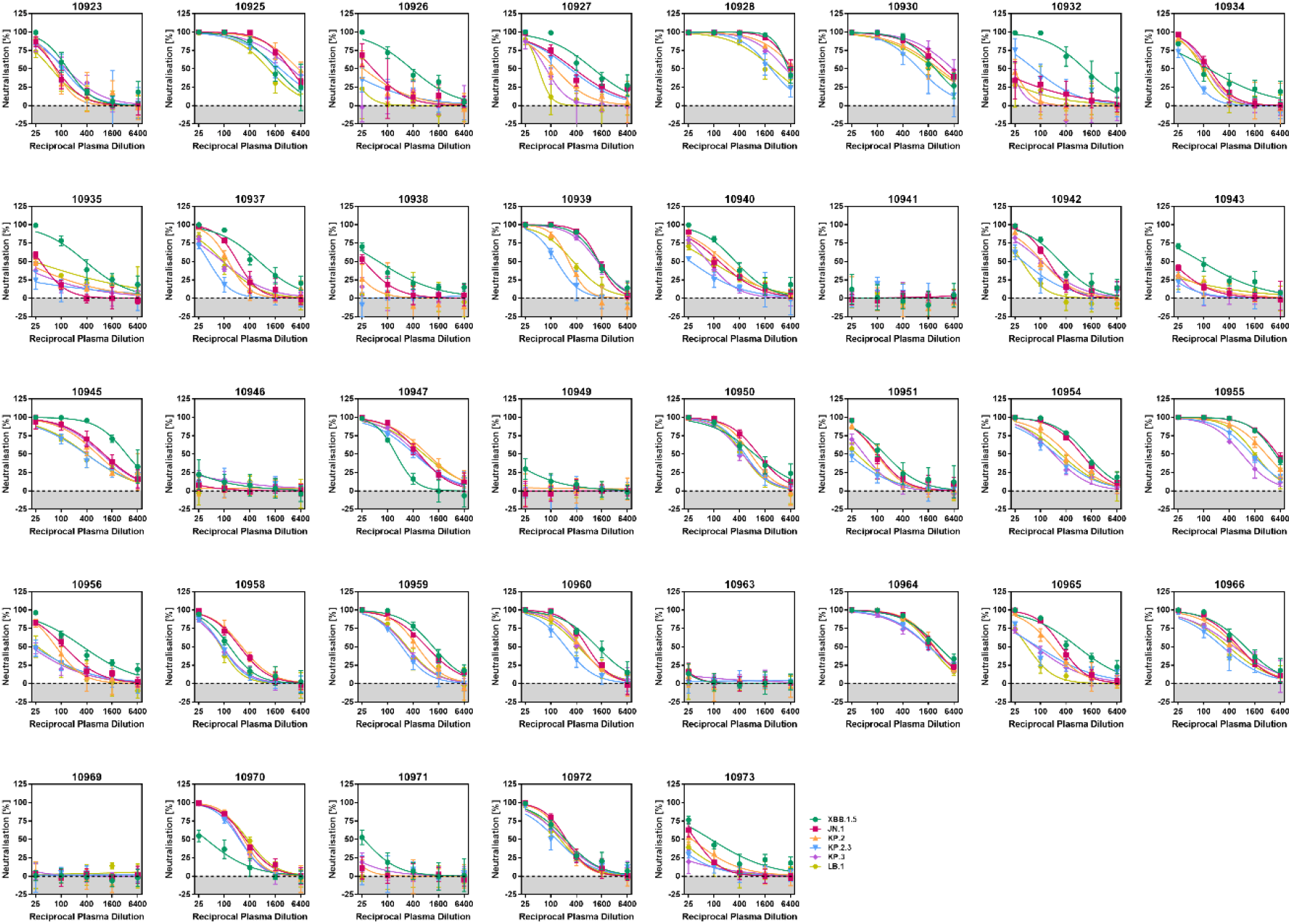
Individual neutralisation data for pre-vaccination plasma.

**Fig. S6.**
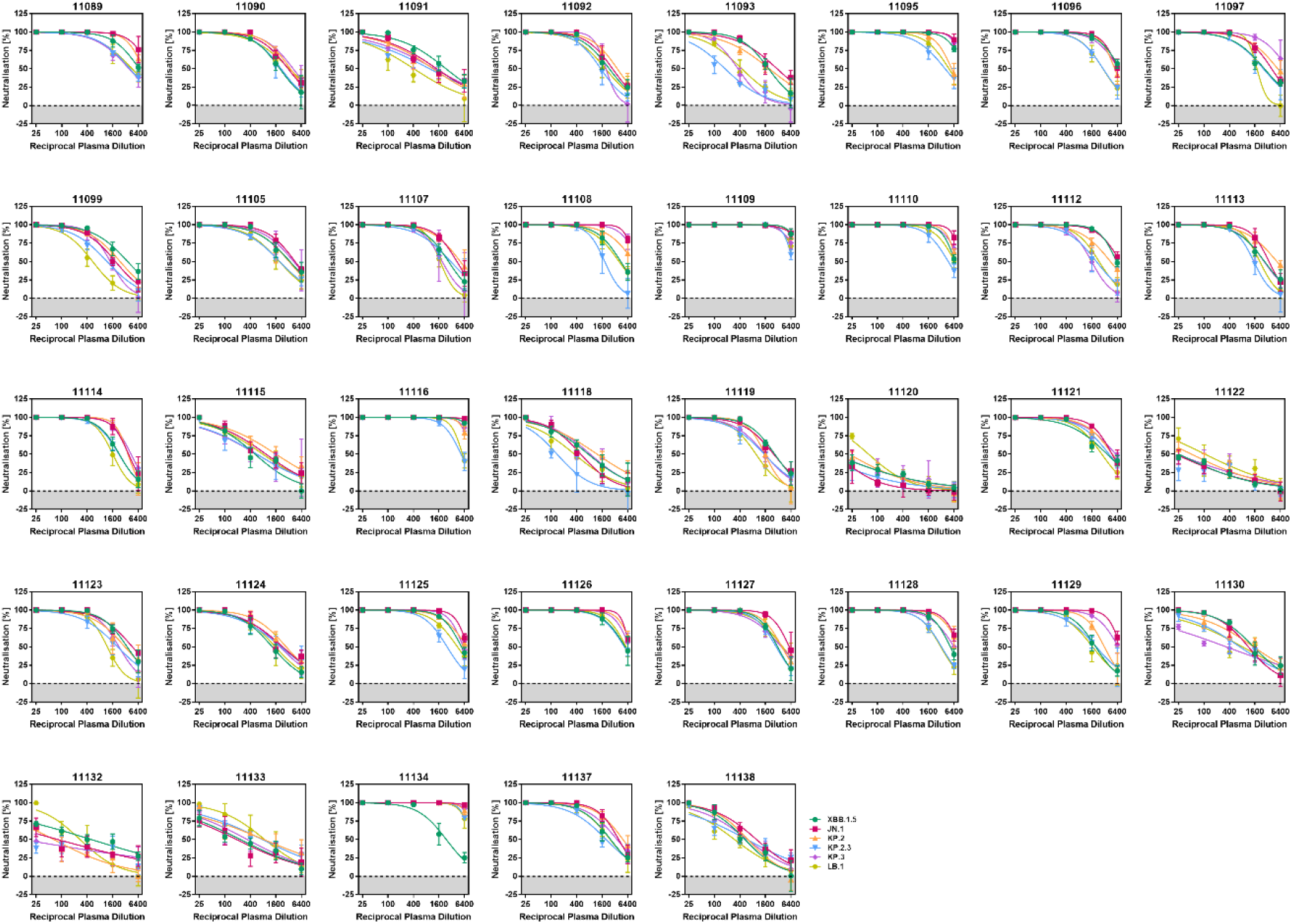
Individual neutralisation data for post-vaccination plasma.

